# Endemic hepatitis B virus (HBV) among hospital in-patients in Bangladesh, including evidence of occult infection

**DOI:** 10.1101/2020.03.17.20037085

**Authors:** Fazle Rabbi Chowdhury, Anna L McNaughton, Mohammad Robed Amin, Lovely Barai, Mili Rani Saha, Tanjila Rahman, Bikash Chandra Das, M Rokibul Hasan, K M Shahidul Islam, M A Faiz, Mamun Al-Mahtab, Jolynne Mokaya, Barbara Kronsteiner, Katie Jeffery, Monique I Andersson, Mariateresa de Cesare, M Azim Ansari, Susanna Dunachie, Philippa C Matthews

## Abstract

Bangladesh is one of the world’s top ten burdened countries for viral hepatitis. We investigated an adult fever cohort (n=201) recruited in Dhaka, to determine the prevalence of hepatitis B virus (HBV) infection and to identify cases of occult hepatitis B infection (OBI). HBV exposure (anti-HBc) was documented in 72/201 (36%), and active HBV infection in 16/201 (8%), among whom 3 were defined as OBI (defined as detectable HBV DNA but negative HBsAg). Applying a target-enrichment sequencing pipeline to samples with HBV DNA >3.0log_10_ IU/ml, we obtained deep whole genome sequences for four cases, identifying genotypes A, C and D. Polymorphisms in the surface gene of the OBI case may account for the negative HBsAg status. We identified mutations associated with nucleos(t)ide analogue resistance, although the clinical significance in this cohort is not known. The high prevalence of HBV in this setting highlights the benefits of offering screening in hospital patients and the importance of HBV DNA testing of transfusion products to reduce the risk of transmission. In order to work towards international Sustainable Development Goal targets for HBV elimination, increased investment is required for diagnosis, treatment and prevention in Bangladesh.

## INTRODUCTION

Global estimates indicate that approximately a third of the world’s population has been exposed to hepatitis B virus (HBV) during their lifetime, with chronic HBV infection (CHB) in over 260 million individuals worldwide ^1^. With an estimated 800,000 annual deaths resulting from CHB, ambitious elimination targets aiming to reduce viral hepatitis incidence by 90% have been established as a part of the UN Sustainable Development Goals for 2030 ^2^. Bangladesh has an intermediate CHB prevalence, estimated at 2-6% ^3,4^, although epidemiology may vary between geographic regions and according to sociodemographic factors ^5^. In combination with the large population, this prevalence rate puts Bangladesh among the top ten high burden countries for viral hepatitis ^6^.

Bangladesh is progressively developing national strategies to tackle HBV ^4^. In 2019, the Government of Bangladesh formulated a ‘National strategic plan for combating viral hepatitis in Bangladesh 2020-2030’ [personal communication, Prof. M. A. Faiz] ^7^. Interventions include programmes for screening donor blood by testing for hepatitis B surface antigen (HBsAg) ^5^, and delivery of universal infant HBV vaccination starting at age six weeks, through the World Health Organization (WHO) Expanded Programme for Immunization (EPI) ^8,9^. Estimates suggest that CHB prevalence has declined from 8% to <6% since the vaccine initiative was introduced in Bangladesh in 2003 ^10,11^, but nevertheless, the perinatal incidence of new infections remains among the highest in South Asia ^12^.

Published HBV sequence data from Bangladesh documents a mixture of genotypes, with genotypes C and D each accounting for ∼40% of the total burden, and genotype A for the remainder, also noting a high proportion of recombinants and mixed genotype infections ^3,5,13^. The phenotype of infection and clinical disease progression may be determined, in part, by these different genotypes ^14^.

Prevalence estimates are typically based on surveillance for HBsAg. However, this does not account for occult HBV infection (OBI), in which individuals are HBV DNA positive, but HBsAg negative (usually in combination with a positive anti-HBc antibody). OBI can arise in several contexts. Most typically, when HBsAg is undetectable, viral loads in the serum are low (<200 IU/ml), reflecting minimal or absent production of HBsAg, or impaired egress of HBsAg from infected cells. A number of mutations have been linked to this low-HBsAg phenotype ^15–17^. HBsAg may also be repressed by anti-HBs seroconversion, and this anti-HBc/anti-HBs occult infection has previously been associated with more severe liver disease ^18–20^. Mutations in HBsAg that affect antibody binding in diagnostic assays have been described, meaning the antigen is expressed but not detected, sometimes referred to as ‘false-occult’ infections ^15^. This is particularly pertinent for assays that rely on detection of the small HBsAg protein, by binding to the ‘a’ determinant. Substitutions, insertions and deletions in this region can therefore result in OBI ^21^. OBI has also been described in the context of co-infection with hepatitis C virus (HCV) ^22,23^, during which HBV replication often appears repressed and HBsAg expression is reduced ^24^. Mechanistic studies have previously suggested that the HCV core protein may directly interfere with HBV replication ^25^.

OBI has been described in Bangladesh amongst blood donors and in individuals with cirrhosis, hepatocellular carcinoma (HCC) and in subjects with raised serum alanine aminotransferase (ALT) ^26,27^. Transmission of OBI can occur through transfusion of blood products that are positive for HBV DNA ^28,29^, and has also been reported from vertical and early-childhood exposure ^30^. There are also documented cases of overt infections transmitting and establishing as transient OBI ^31,32^. The majority of clinical and epidemiological studies do not detect OBI, as screening based on HBV DNA is expensive, and a large number of HBsAg-negative individuals have to be tested to identify those who are HBV DNA positive, particularly if the overall population prevalence is low. For this reason, the true burden and impact of OBI is difficult to assess. However, recommendations for screening blood products include the need for HBV DNA testing, rather than reliance on HBsAg screening alone, in order to avoid transmission events as a result of OBI ^33^.

Recognising the high burden of HBV infection in Bangladesh, and limited data about OBI, we set out to investigate the prevalence and molecular characteristics of HBV, and to determine whether cases of OBI were present, using samples obtained from a cohort of adult hospital in-patients.

## METHODS

### Study settings and clinical cohort

We used the opportunity of a hospital cohort recruited in Bangladesh to study the epidemiology of adult HBV infection in this setting, anticipating that HBV infection would not be relevant to the diagnosis for which they had presented to hospital. Serum samples were collected from patients enrolled into a prospective observational clinical fever cohort recruited at two sites:

i. Dhaka Medical College Hospital (DMCH) is a public hospital, and the largest tertiary care hospital in Bangladesh with 2300 beds. Patients are admitted to a full range of specialties, and mostly receive free care.
ii. The Bangladesh Institute of Research and Rehabilitation in Diabetes, Endocrine and Metabolic Disorders (BIRDEM), is a non-government not for profit specialist hospital in Dhaka with 750 beds. BIRDEM receives regular annual subsidy from the Bangladesh ministry of social welfare and other sources. Baseline investigations are performed at a government-subsidised rate, but in the majority of cases patients need to pay for more specialist investigations.

Both hospitals receive patients from all over the country, and are therefore considered tertiary referral centres. This cohort recruited adults (≥18 years of age) with a history of fever >38°C for >48 hours from Internal Medicine, Critical Care Medicine, Surgery and Orthopaedics between June-October 2017. As routine infant HBV vaccination was not rolled out in Bangladesh until 2003, few of this cohort are likely to have received childhood immunisation against HBV.

### Ethics

Ethical approval was obtained from the Research Ethics Committee (REC) of DMC (MEU-DMC/ECC/2015/97), BIRDEM (BADAS-ERC/EC/17/0127) and Oxford Tropical Medicine Research Ethics Committee (OXTREC: 51-16) before the commencement of the study. Participants provided written informed consent for participation.

### Blood sampling and baseline screening for HBV infection

Serum samples were collected and screened for HBV infection and HBV exposure at local accredited testing facilities (Virology department of Bangabandhu Sheikh Mujib Medical University) in Dhaka, using HBsAg (LIAISON® XL Murex HBsAg Quant, Italy) and total anti-HBc (LIAISON® Anti HBc, Italy) respectively. All HBsAg-positive and anti-HBc-positive patients were screened for HBV DNA at the clinical diagnostic laboratory at Oxford University Hospitals NHS Foundation Trust, UK (COBAS AmpliPrep/COBAS TaqMan, Roche, Welwyn Garden City, UK ^34^), an automated platform to detect and quantify HBV DNA).

### Clinical follow-up

Patients who tested HBV positive were informed about the result and referred to the Hepatology department of the respective institutes for appropriate clinical assessment, management and follow-up.

### DNA extraction and sequencing

Samples with HBV DNA viral load ≥3.0 log_10_ IU/ml underwent a target-enrichment approach for HBV whole genome sequencing (WGS) on an Illumina Mi-Seq platform in Oxford, UK. Total nucleic acid was extracted from patient serum using the NucliSENS magnetic extraction system (bioMérieux). A completion-ligation reaction was performed to convert the partially dsDNA genome into a fully dsDNA molecule ^35^, after which nucleic acid was purified using Agencourt RNAClean XP magnetic beads (Beckman Coulter).

Sequencing libraries were generated using the NEBNext Ultra II DNA Library Prep Kit (New England Biolabs) and libraries were assessed using TapeStation system (Agilent) and Qubit dsDNA HS Assay (Thermo Fisher Scientific). An adapted target-enrichment workflow was applied to the SeqCap EZ (Roche) protocol, using custom-designed HBV probes ordered from IDT (xGen Lockdown Probes). Samples were sequenced on an Illumina Mi-Seq using a v3 300bp paired end kit.

### Analysis of sequence data

Deep sequencing read pairs were de-multiplexed using QUASR v7.01 and adapter sequences trimmed with CutAdapt v1.7.1 ^36^. Short reads (<50bp length) and reads mapping to the human genome reference sequence (identified using Bowtie v2.2.4 ^37^) were discarded. Remaining reads were mapped to HBV reference sequences representing genotypes A-I using BWA mem v0.7.10 ^38^, to select the most appropriate reference sequence and to identify HBV reads. Reads were then mapped against the reference sequence using BWA mem and consensus sequences were derived. Simmonics Sequence Editor ^39^ was used for alignment and sequence examination, with the genotype A sequence X02763 used as a numbering reference. Maximum-likelihood phylogenetic analysis of consensus sequences was performed with MEGA7 ^40^ with 1,000 bootstrap replicates used. Trees were visualised in Figtree ^41^. Sequences were analysed with reference sequences ^42^ and 61 published full length HBV genome sequences from Bangladesh identified in Genbank.

Sequences were checked for evidence of recombination by bootscan analysis using RDP4 ^43^.

### Identification of potential resistance associated mutations (RAMs) and vaccine escape mutations (VEMs)

We used previously published lists of HBV drug resistance mutations for lamivudine (3TC) ^44^ and tenofovir disoproxil fumarate (TDF) ^45,46^ as a reference against which to check our sequence data for the presence of specific RAMs. VEMs are most commonly reported within the HBV surface antigen (HBsAg) ‘a’ determinant (residues 124-147), which is the target of neutralising antibodies. We therefore focused scrutiny on this region, searching for evidence of polymorphisms Q129H/R, M133L, F/Y134I/L/T, K141E, P142S and G145R/A that have been highlighted by a recent paper taking a systematic approach to the identification of VEMs in global data ^47^. We also added A128V to this list, as this has been independently reported as a VEM in Bangladesh ^48,49^.

### Statistics

Data were analysed using the software package GraphPad Prism version 7.0 (San Diego, CA, USA). Two-sided p values were calculated using the Chi-square and Fisher’s exact test for dichotomous and ordinal variables. Continuous variables were compared using one-way ANOVA and the Mann-Whitney U-test. Demographic factors and clinical characteristics were summarized with counts (%) for categorical variables and median (interquartile range [IQR]) and mean (±standard deviation) for continuous variables.

## RESULTS

### Cohort description and HBV epidemiology

The cohort recruited 201 adults, with a median age of 35 years (IQR 28-55 years), of whom 131/201 (65%) were male. Patients travelled a median of 100 km (IQR 20-165km) to the hospital, with a proportion of patients (28/201, 14%) travelling >300km, reflecting the status of these hospitals as specialist centres.

Exposure to HBV in this cohort was common, with 72/201 (35.8%) samples testing anti-HBc-positive. Active HBV infection was confirmed in a total of 16/201 individuals (7.9%). These comprised ten who were positive for both HBsAg and HBV DNA (10/201, 4.9%), three positive for HBsAg with undetectable HBV DNA (3/201, 1.5%), and three with OBI based on testing positive for HBV DNA with negative HBsAg (3/201, 1.5%) (Fig 1). Median viral load of the samples with detectable HBV DNA was 2.6 log_10_ IU/ml (IQR 1.9-5.0 log_10_ IU/ml). One overt HBV infection was excluded when estimating the median viral load as the sample was reported as HBV DNA positive, but with a viral load below the limit of quantification (<1.0 IU/ml).

**Figure 1:**
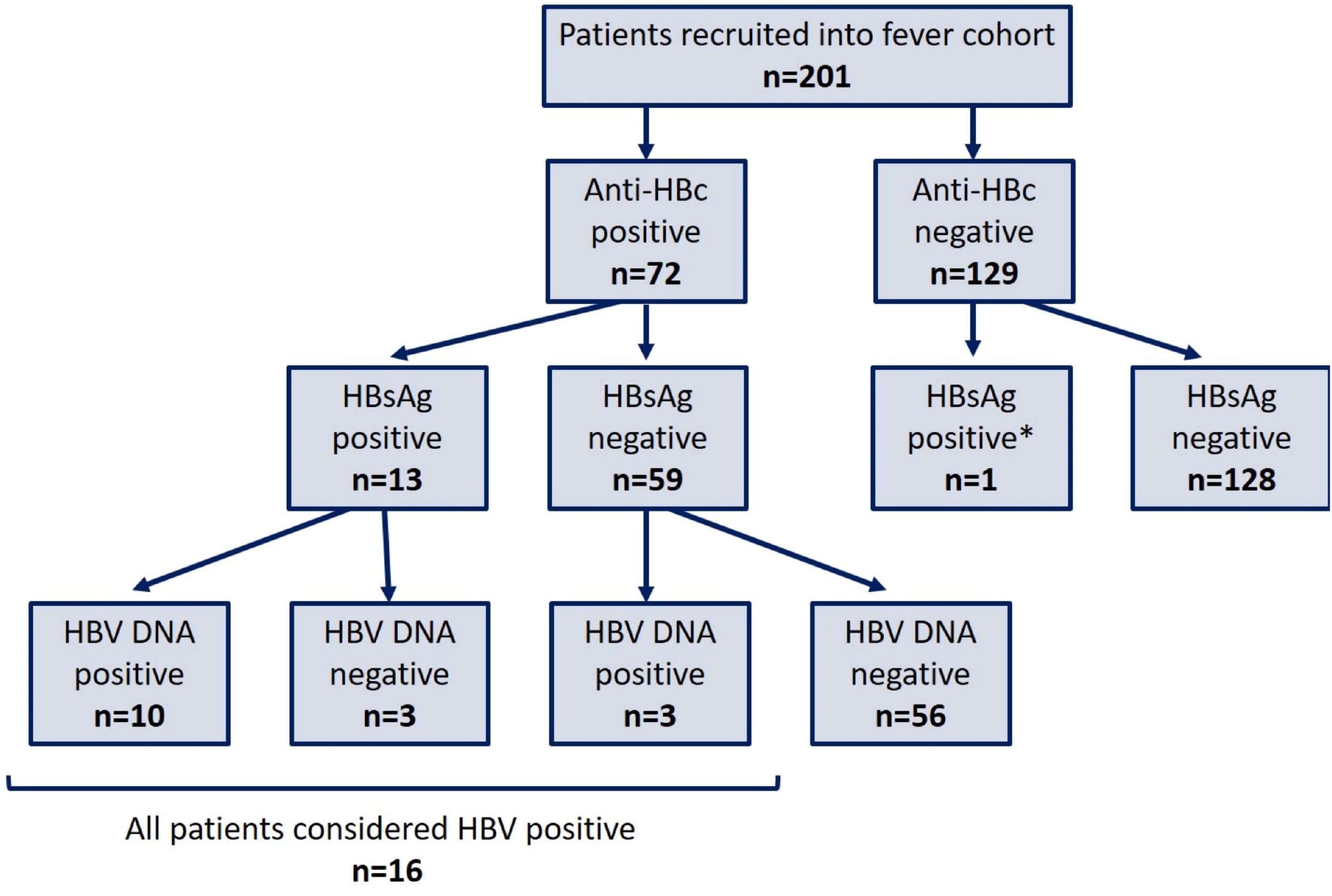
Flow chart illustrating the results of screening for HBV markers in serum from adults in a Bangladesh fever cohort. All patients were screened for both anti-HBc and HBsAg. All HBsAg positive patients were further screened for HBV DNA. A total of 16 patients were HBV positive, with three different serological profiles; (a) HBsAg positive, HBV DNA positive; (b) HBsAg positive, HBV DNA negative; (c) HBsAg negative, HBV DNA positive (occult HBV infection). *One patient was anti-HBc negative, HBsAg positive and HBV DNA negative. This patient was likely a false reactive on the HBsAg screening test and considered to be HBV negative for the study.

### Characteristics of HBV infection

There was no significant difference in sex or age between individuals with or without HBV infection (p=0.18 and p=0.15, respectively; Table 1). Patient occupation was recorded, with HBV exposure being more common amongst homemakers (p=<0.001) and relatively infrequent in students (p=0.02) (Table 1). The median duration between symptoms and hospitalization was 10 days across all groups (p=0.18) (Table 1). There was no significant difference in patient outcome following their admission with fever according to HBV infection status (p=0.81). We detected OBI only in male patients ≥60 yrs of age, which is significantly older than the age of the other HBV-positive subjects (Fig 2A, p=0.02). Median HBV viral loads of the samples with occult infection (2.6 log_10_ IU/ml, IQR 1.1-5.3 log_10_ IU/ml) were comparable to the viral loads of the HBsAg-positive individuals (2.4 log_10_ IU/ml, IQR 1.8-4.7 log_10_ IU/ml) (Fig 2B, p=0.94). One of the OBI infections (sample ID 118) presented with a viral load of 5.3 log_10_ IU/ml, which is unusually high for occult infection, and indicates ongoing high levels of viral replication.

**Table 1.** Characteristics of patients recruited into the study cohort at two teaching hospital centres in Dhaka, Bangladesh.

|  | HBV positive<br>(HBsAg or HBV<br>DNA positive) | Resolved HBV<br>(anti-HBc<br>positive only) | HBV negative<br>(anti-HBc<br>negative) | p-value<sup>a</sup> |
| --- | --- | --- | --- | --- |
| **Number of patients**<br>**(%)** | 16 (7.9%) | 56 (27.9%) | 129 (64.2%) | - |
| **Median age**<br>**in yrs (IQR)** | 40 (30-58) | 38 (28-60) | 34 (21-55) | 0.15 |
| **Male sex, *n* (%)** | 10/16 (62.5%) | 39/756 (69.6%) | 72/129 (55.8%) | 0.18 |
| **Median distance in**<br>**km travelled to**<br>**hospital (IQR)** | 135 (28-150) | 70 (16-200) | 100 (28-150) | 0.81 |
| **Median number of**<br>**days between**<br>**symptom onset**<br>**and hospitalization**<br>**(IQR)** | 10 (7-12) | 10 (6-15.8) | 10 (6-15) | 0.18 |
| **Occupation *n* (%)** |  |  |  |  |
| **Homemaker** | 4 (25.0%) | 20 (35.7%) | 19 (14.7%) | <0.001 |
| **Student** | 1 (6.3%) | 10 (17.9%) | 41 (31.7%) | 0.02 |
| **Farmer / day**<br>**labourer** | 8 (50%) | 22 (39.3%) | 43 (33.3%) | 0.37 |
| **Others<sup>b</sup>** | 3 (18.7%) | 4 (7.1%) | 26 (20.1%) | 0.09 |
| **Outcome *n* (%)** |  |  |  |  |
| **Died in hospital<sup>c</sup>** | 3 (18.8%) | 8 (14.3%) | 34 (26.3%) | 0.18 |

<sup>a</sup> One-way Anova and Fisher's exact test (two tailed) was applied
<sup>b</sup> Includes businessman, carpenter, plumber and unemployed
<sup>c</sup> All other patients were either discharged after (n=154) or they absconded during (n=2) treatment in hospital

**Figure 2:**
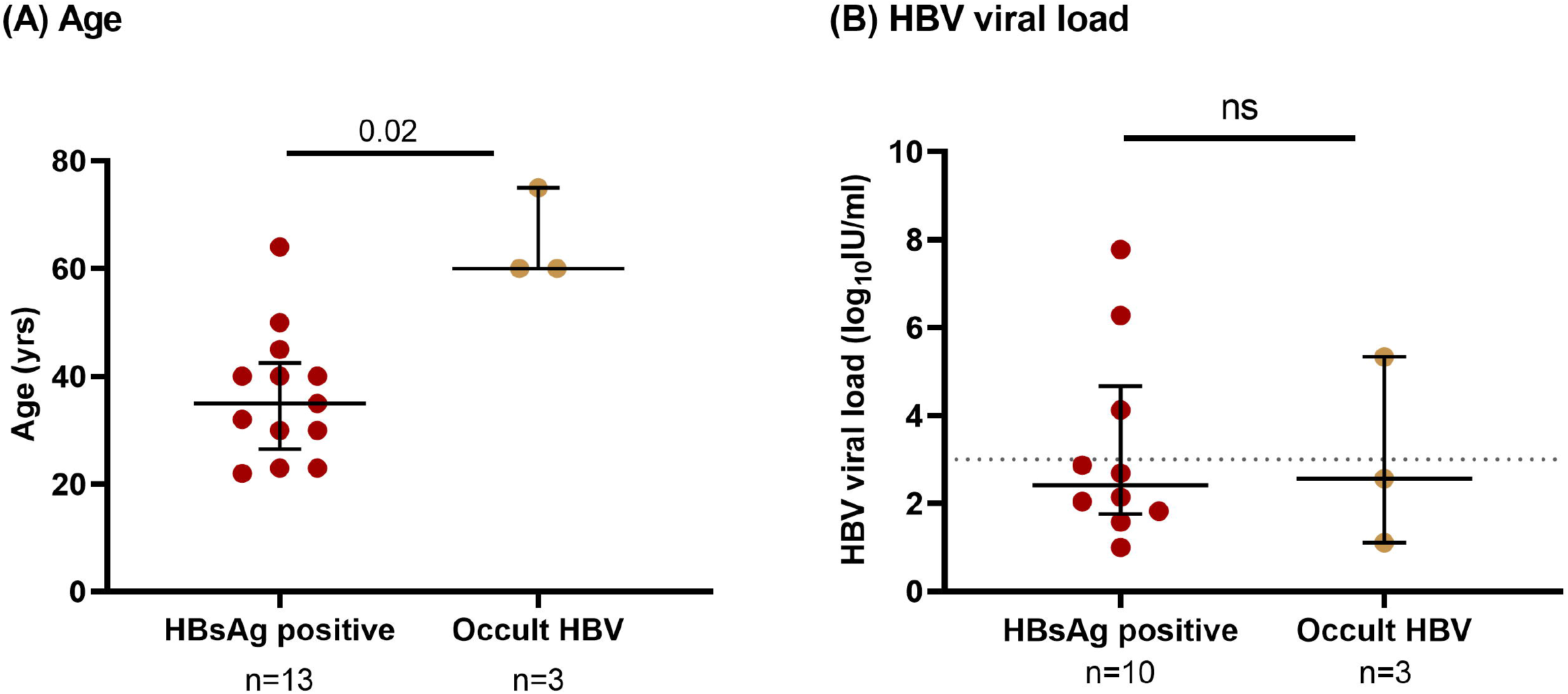
Characteristics of adults in Bangladesh with HBV infection, comparing those with HBsAg-positive infection to those with occult HBV infection. (A) Ages and (B) viral load of patients with HBsAg-positive HBV and occult HBV infections. Three samples were HBsAg-positive. Median and interquartile range (IQR) are indicated for both graphs in blue. The sensitivity threshold for deep sequencing (3.0log_10_ IU/ml) is indicated in figure B with a dashed red line.

### Identification of HBV genotypes A, C and D

We obtained full length HBV genome sequences from serum from all four individuals with HBV viral loads ≥3.0 log_10_ IU/ml, of whom one had OBI (ID 118). Median coverage across the genome ranged from 33-77,644 reads per site. Low viral loads and reduced sample volumes were associated with reduced coverage (Fig 3). The consensus sequence for sample 197 indicated a truncated HBeAg protein, based on a G1986A mutation present in 57% reads. This trunation is well-described in HBV, and frequently linked to HBeAg seroconversion ^50^. Both 051 and 118 also expressed the truncated HBeAg protein as a minority variant, present in 32% of reads in both samples. Phylogenetic analysis indicated two genotype C infections, and one each of genotypes A and D (Fig 4). The OBI sequence grouped with other genotype C sequences. Based on the phylogeny of full length sequences and bootscan analysis, we did not find evidence of recombination in this dataset.

**Figure 3:**
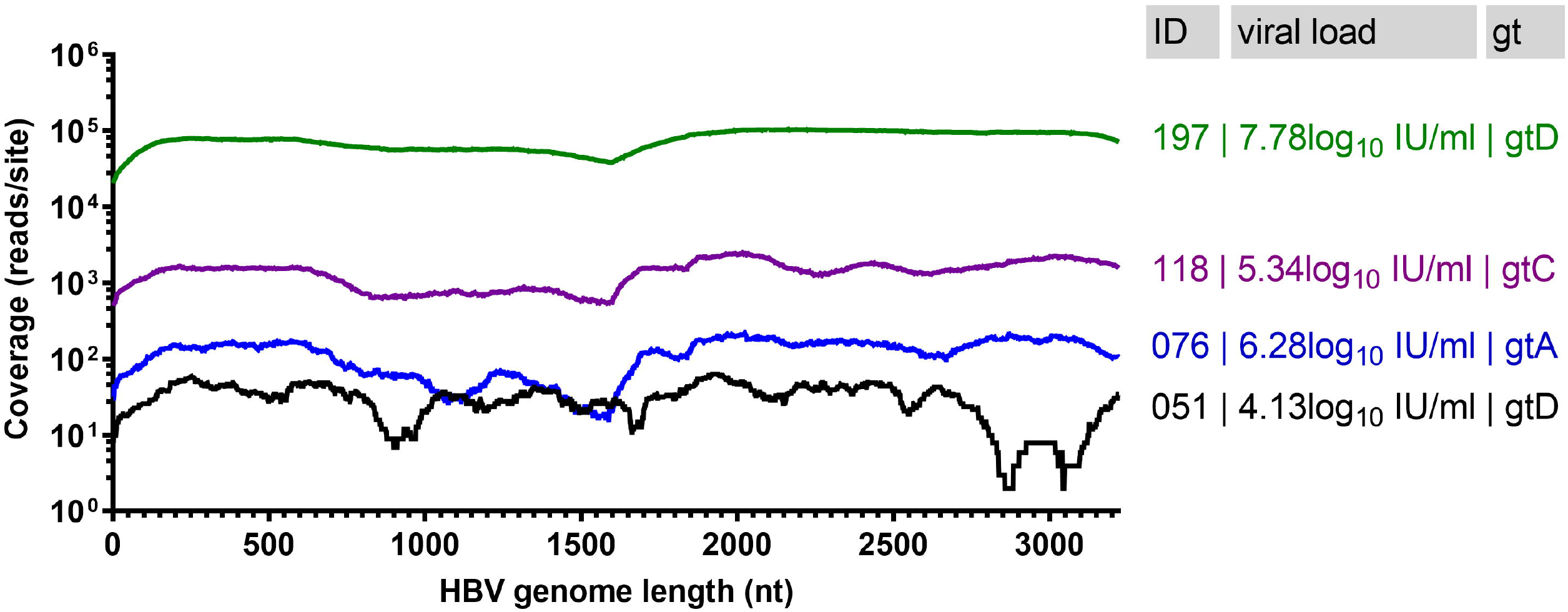
Plot showing full length coverage of the HBV genome based on sequencing by Illumina for four serum samples from adults in Bangladesh. Sample IDs, viral loads and viral genotype (gt) are given for each sequence. Sample ID 118 was a case of occult HBV infection.

**Figure 4:**
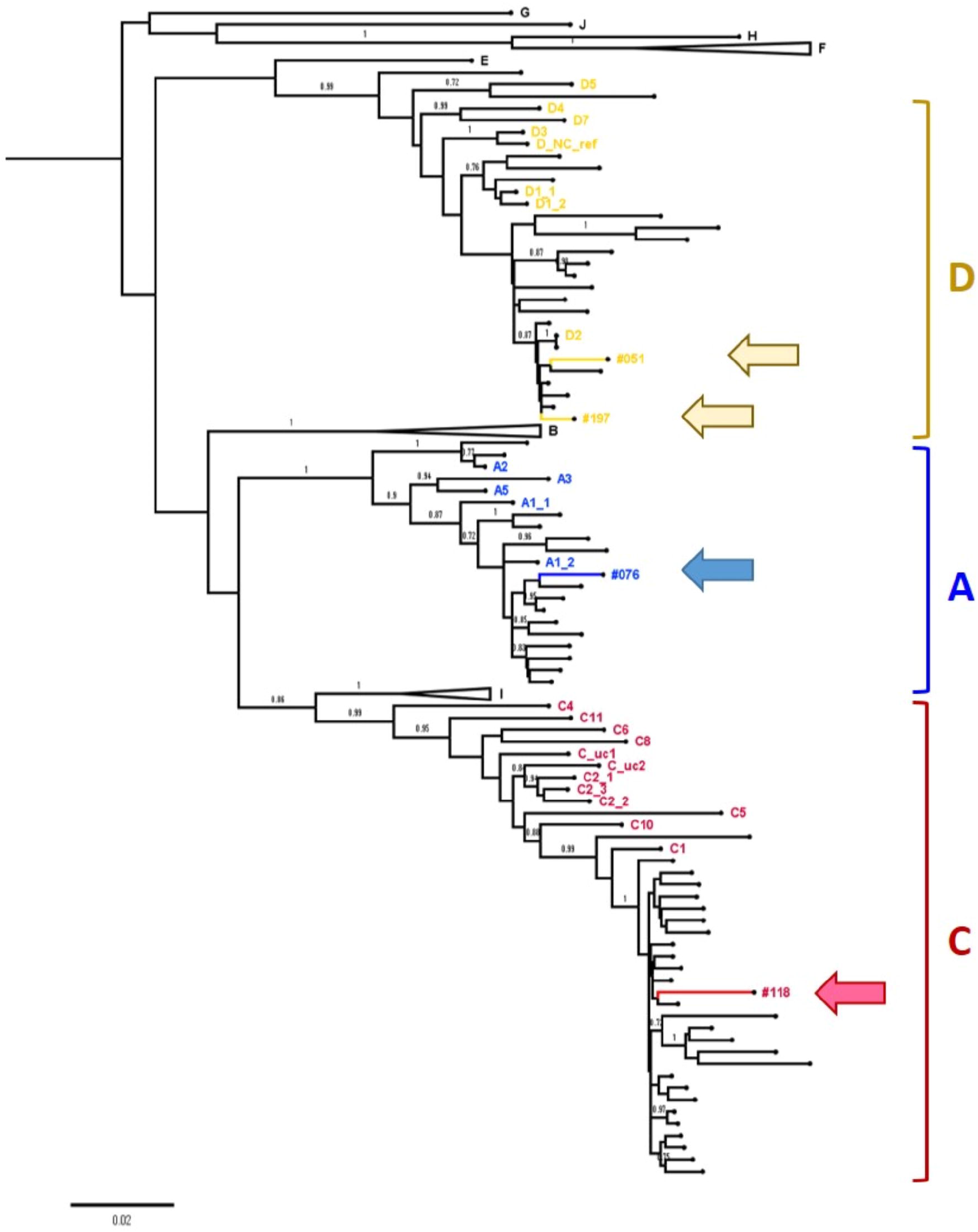
Phylogenetic tree to show the relationship between new HBV sequences and published HBV sequences from Bangladesh. Four full-length HBV consensus sequences generated in this study were analysed alongside HBV genotype reference sequences (for genotypes A-J) ^42^ and 61 sequences originating from Bangladesh identified in Genbank ^66^ (unlabelled branches, Suppl Table 1). Genotype A sequences are highlighted in blue, genotype C in red and genotype D in yellow. The four sequences generated in this study are indicated with arrows; we identified one genotype C1 sequence (sample 118), one genotype A1 (sample 076) and two genotype D2 sequences (051 and 197). Genotype clades without Bangladesh sequences clustered in them have been collapsed. Bootstrap replicates were repeated 1000 times, and all branches with support >70% are indicated.

#### HBV Resistance Associated Mutations (RAMs)

We examined the consensus reverse transcriptase (RT) sequences for the presence of RAMs and VEMs (Table 2). Polymorphisms A181T and Q215P in sample 197 have been associated with resistance to 3TC, and may be sufficient to cause a drug resistant phenotype ^45^. The overlapping reading frames in HBV mean the A181T mutation corresponds to the HBsAg W172* mutation, which has been associated with progressive liver disease ^51^.

**Table 2:**
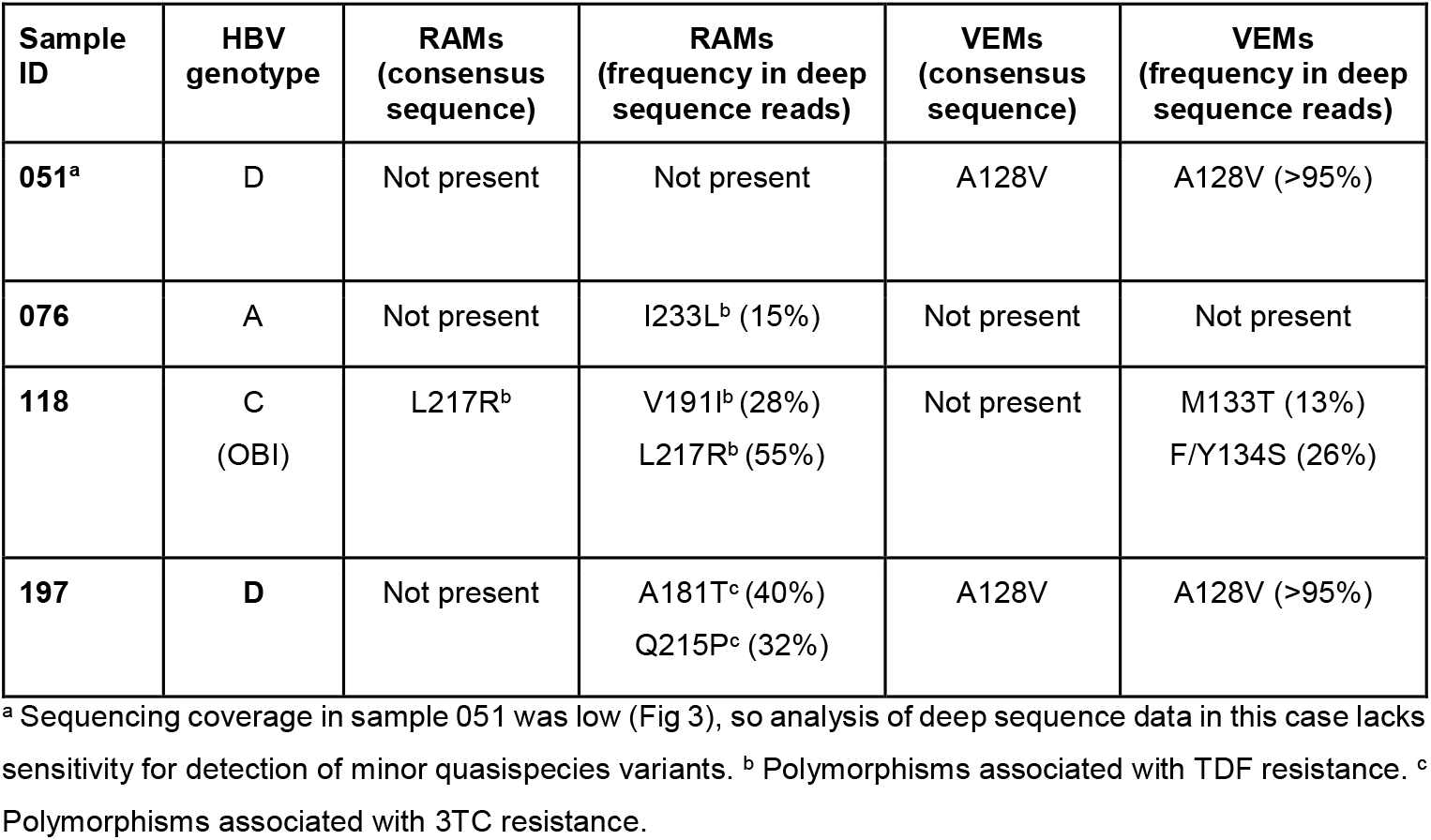
Presence of Putative Resistance Associated Mutations (RAMs) and Vaccine Escape Mutations (VEMs) in samples from four adults with chronic HBV infection (including one occult infection, OBI), based on analysis of consensus level data and deep sequences generated by Illumina. Positions for codons associated with RAMs are listed in RT, and for VEMs are in HBsAg (sites all numbered using genotype A sequence X02763 as a numbering reference).

There is currently a lack of robust evidence for TDF resistance in HBV, but emerging reports to suggest that certain combinations of RT polymorphisms may be associated with reduced susceptibility *in vivo* ^*46*^. We interrogated our sequences for any of these polymorphisms (Table 2). L217R was present at consensus level in sample 118 (55% of reads) and V191I was identified as a minority variant (28% reads). I233L was present as a minority variant (in 15% of reads) in sample 076. The clinical significance of these substitutions is not well understood, but phenotypic resistance has most robustly been described when in combination with ≥2 other RT RAMs ^46 44^.

VEMs M133T and and F/Y134S were present as minority variants in sample 118. A128V has been highlighted as a putative VEM based on previously reported sequence data from Asia, and was identified at consensus level in both our genotype D sequences. However based on subgenotype (D2 for both 051 and 197), valine is consensus at this position which makes its provenance and clinical significance unclear.

#### Investigation of sequence data from patient with OBI

We examined the OBI sample (ID 118) for the presence of polymorphisms previously linked to OBI, based on previously published catalogues of relevant sequence changes ^52,53^. There were no HBsAg (pre-S1, pre-S2 and S) OBI-associated variants in the consensus sequence. Examining the deep sequencing reads for the surface gene we identified an 18bp deletion in 10% of reads at the start of pre-S1, with a further 20% of reads having W4R/S5Y mutations and W4P identified in 5% of reads (Fig 5). These polymorphisms have been previously associated with reduced HBsAg expression ^52,53^. I84T/M was identified in 48% of reads, and again appears more common in individuals with occult infection ^52^. An ACG mutation was present in place of the start codon (ATG) in 7% of reads (mutation also referred to M1T), suggesting a number of isolates were incapable of producing pre-S2. A deletion was also present at aa30/31 in pre-S2, although it is unclear if this deletion has any association with OBI. In the S region, 28% of reads had a W182* mutation, generating a truncated HBsAg product, which is also associated with OBI ^52^. The W182* mutation is mirrored in RT as V191I, which has been linked to TDF resistance (Table 2). The ‘a’ determinant of the virus had minority variants that have been previously associated with vaccine escape (M133T and F/Y134S), but their provenance in OBI remains unclear.

**Figure 5:**
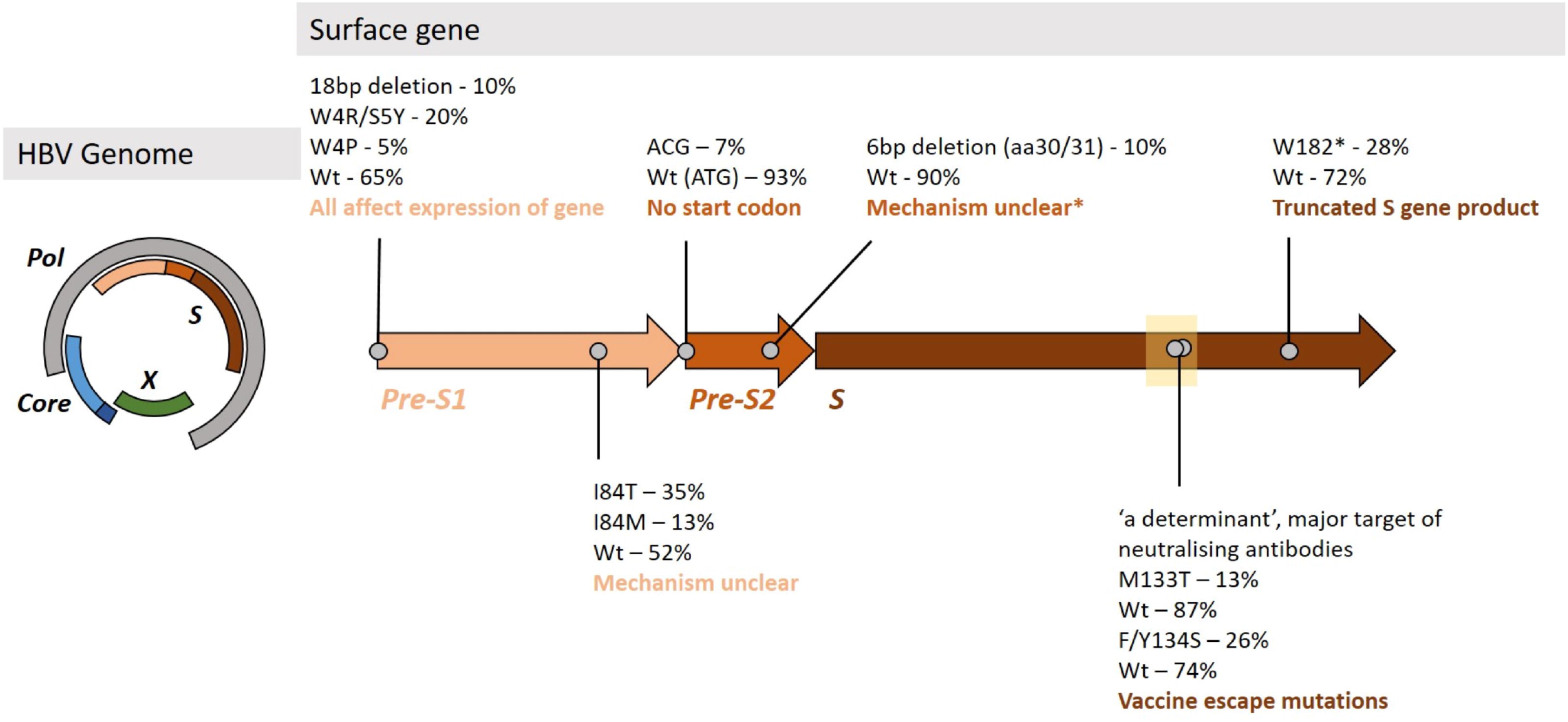
Mutations identified in the Surface gene of sample 118 that may be linked with the OBI phenotype. The HBV surface gene is subdivided into three domains, pre-S1, pre-S2 and S. The proportion of reads containing polymorphisms are shown at various positions throughout the gene. Wt - wild type, aa - amino acid. The ‘a’ determinant (marked with a box at residues 124-147), is a major target of neutralising antibodies and widely-used target for diagnostic assays, and has a strong association with OBI mutations ^17,21,53,67^. *The deletion identified at aa30/31 in pre-S2 has not been associated with OBI previously, and it is unclear if this deletion is associated with OBI as the mechanism inducing OBI is unclear.

## DISCUSSION

Our study confirms the presence of HBV infection in Bangladeshi adults, with an overall prevalence of 8% in an urban hospital cohort, including a contribution made by OBI. Despite the size of our cohort and the limitations regarding which samples could be sequenced, we identified a range of genotypes, in keeping with previous data from Bangladesh ^5,13^. Of note was the identification of a genotype A1 isolate, more typically associated with transmission in Southern and Eastern Africa. However, sequences belonging to this subtype have been reported previously in the region ^3^, and may have increased in frequency over the past decade ^5^.

### Epidemiology of infection

Our cohort is too small to make any generalisations about the distribution of HBV infection in this setting. However, we noted students were significantly less likely to be exposed than other adults, which may reflect younger age and better socioeconomic backgrounds, both of which could make exposure less likely and vaccination more likely. Those with ‘homemaker’ status were significantly more exposed; this may be associated with gender or socio-economic status.

### Evidence for drug resistance

A previous meta-analysis of drug resistance in Bangladesh reported a prevalence of 3TC resistance of 11%, but no TDF resistant motifs were described ^48^. Although our cohort is too small to draw any further conclusions about the prevalence of RAMs and VEMs in this population, it is striking that - even in a small number of sequences - we identified mutations that have been highlighted in reports of resistance to both 3TC and TDF (Table 2). In these treatment-naive patients we cannot confirm the *in vivo* significance of any of the RAMs we have reported and there is currently insufficient evidence to correlate TDF RAMs robustly with a drug resistant phenotype *in vivo*. However, our data do raise concern that these mutations are circulating in this setting; further work is urgently required to explore the overall prevalence and clinical significance of these variants in individual patients and at population level in Bangladesh.

### Evidence for vaccine resistance

Mutations in HBsAg at positions 133 and 134 (arising in sample 118) are localised within the major hydrophilic region of the surface protein (‘a’ determinant, residues 124-147) known to contain major B-cell epitopes and can therefore contribute to vaccine resistance ^45,54,55^. A128V is reported in sequence surveillance data representing China in the post-vaccine era, where it is highlighted as a potential VEM, although its specific contribution to vaccine resistance remains uncertain ^56^. Previous reports of VEMs in Bangladesh ^48^, and a case report of a child, ^49^ have also identified the A128V substitution, and we identified this variant in two of our sequences (both subgenotype D2). However, it should be noted that a recently proposed subgenotype D2 reference sequence (MF925358) ^42^ has 128-valine, suggesting it is the most common residue in this subgenotype. This raises the possibility that subgenotype D2 might be more susceptible to vaccine resistance, but further work is required to substantiate this. The significance of these variants in our sequence data is uncertain, as we did not collect vaccine history data or measure vaccine-mediated antibody titres.

### Polymorphisms associated with OBI

Sequencing data from a case of OBI demonstrated a combination of mutations and deletions, occurring in all three regions of the surface gene. None of the deletions were present at consensus level, illustrating how the application of deep sequencing allowed us to identify variants that would not have been visible at consensus level. Several of the mutations identified have been linked with the progression of severe liver disease, including W182*, thought to interfere with cell cycle regulatory processes via inhibiting TGFB-1 expression ^57^ and linked to severe liver disease even in the setting of low viral loads ^51^.

### Implications for policy, clinical practice and resources

It is likely that HBV infection was incidental to the presenting syndrome in all of these individuals, being a bystander rather than the index cause of illness requiring hospital admission, as evidenced by the similar outcomes for admission with fever among HBV-negative and -positive patients. The approach we adopted here demonstrates the feasibility of adding routine HBV screening into investigations for adults admitted to hospital, which is one potential approach to systematic enhancement of HBV diagnosis. At a population level, improvements in ascertainment will be of crucial importance in advancing progress towards international elimination goals ^6^. Diagnosis also provides individual benefits; in this case, we provided HBV diagnoses that would have otherwise gone undetected, facilitating referral into appropriate clinical care and follow-up for individuals.

We have documented a persistent ongoing burden of HBV infection in adults, which highlights the importance of continued deployment of vaccination programmes with a focus on infant coverage to reduce vertical and early-life transmission events. In other high/medium endemicity settings, there has been strong advocacy to introduce birth dose vaccination in order to further reduce the incidence of mother-to-child transmission ^58^. Screening blood products for occult HBV infection using HBV DNA rather than HBsAg should be considered where possible to prevent transmission in cases of OBI ^59^. The cost-effectiveness of such additional screening remains to be formally assessed in Bangladesh, but costs can be reduced by pooling sera for screening, and only testing individual samples from within pools that test positive ^60^. Combination anti-HBc and ALT screening has also been suggested as a lower-cost alternative ^27^.

There are resource implications for any approach to upscaling HBV diagnostics, that include not only the cost of the screening tests, but further characterisation of infection to stratify patients for therapy, including HBV DNA viral loads and/or HBeAg. Costs are also incurred through the need for further clinical assessment and investigations. However, simplified screening approaches have been proposed for use in resource-constrained settings ^61^, and generic, locally-produced, first line antiviral drugs for HBV infection, tenofovir and entecavir, are available in Bangladesh ^4^.

### Caveats and limitations

We have undertaken HBV screening on only a small cohort. Although HBV was presumed to be incidental to the reasons for presentation to hospital, we cannot exclude a selection bias that enriched for HBV infection. We have assumed that HBV infections were likely to be chronic, but have not distinguished between anti-HBc IgM and IgG, so cannot exclude the possibility of acute HBV infection. Although the population represented a wide geographic area, we have captured only a very limited cross-section of the overall population, without any longitudinal follow-up. Subjects in the cohort were tested predominantly for biomarkers relating to sepsis, and routine blood tests used to assess HBV infection were not available (and interpretation would be confounded by infection). Subjects were not routinely screened for HCV, which has been associated with occult HBV infection and is moderately prevalent in the region, with approximately 1% of the population infected ^6,10^.

The study highlights the current technical challenges of generating full length HBV sequence data from cases of HBV infection, with a threshold HBV DNA of >3.0log_10_ IU/ml typically required ^62,63^. The median viral load of 2.6 log_10_ IU/ml in this cohort illustrates the high proportion of cases with viral loads below the current sensitivity of the sequencing platform ^63,64^. For this reason, we generated full length sequence data in only 4/13 cases, leaving the majority of HBV sequences ‘under the radar’, including two OBI. The lack of highly sensitive sequencing methods for HBV remains an ongoing issue in furthering our understanding of HBV disease and transmission.

Short read sequencing technology provides advantages in sequencing depth, but reconstruction of full length haplotypes is challenging, making it difficult to determine whether there is linkage between mutations in the sequences. Given that no mutations linked to OBI were identified at the consensus level in sample 118, this suggests that all sequences must carry at least one of the mutations observed in the HBV surface gene, and reliance on short-read sequencing technology is limiting our current understanding of how these mutations interact to produce a particular phenotype. Many different mutations have been described in OBI, and the individual or combined influence of each of these is often not well defined; furthermore, relevant polymorphisms may differ between genotypes ^65^.

## Conclusions

Given the high burden of HBV infection in Bangladesh, there is an urgent need to expand our understanding of epidemiology, including OBI, in order to strengthen advocacy for interventions that include diagnostic programmes, enhanced screening of blood products using nucleic acid testing, and birth dose infant vaccine. In the decade ahead, investment in clinical care and public health interventions, education and research will be crucial to underpin advances towards international elimination targets for Bangladesh.

## Data Availability

The datasets generated during and/or analysed during the current study are available in the following repositories:
(1) GenBank accession numbers MT114170 - MT114173
(2) Metadata file of demographic data and HBV screening results in the fever cohort (n=201 subjects) on Figshare - 10.6084/m9.figshare.11973930
(3) A STROBE statement has been submitted as a supplementary file with this manuscript.

DOI:10.6084/m9.figshare.11973930

## AUTHOR CONTRIBUTIONS

FRC, ALM, SD and PCM conceived and designed the project. FRC and SD sought ethical approval for the study. BCD and MRH recruited patients and obtained informed consent for the testing. Blood samples were tested by LB, MRS, TR, KJ and MIA. Sample preparation and Illumina sequencing was carried out by AM and MdC, with bioinformatic analysis from MAA. Data analysed by FRC, ALM, and JM. FRC, ALM and PCM wrote the manuscript with input from KMSI, MAF, MA-M, MRA, MAA, BK and SD. All authors read and approved the final manuscript.

## DATA AVAILABILITY

The datasets generated during and/or analysed during the current study are available in the following repositories:

- GenBank accession numbers MT114170 - MT114173
- Metadata file of demographic data and HBV screening results in the fever cohort (n=201 subjects) on Figshare - 10.6084/m9.figshare.11973930
- A STROBE statement has been submitted as a supplementary file with this manuscript.

## SUPPLEMENTARY DATA FILE

**Suppl Table 1: Accession numbers of full-length HBV sequences identified in GenBank originating from Bangladesh**. The sequences were analysed alongside the four consensus sequences generated from our study (Fig 3).

